# ARB/ACEI use and severe COVID-19: a nationwide case-control study

**DOI:** 10.1101/2020.06.12.20129916

**Authors:** Hee Kyoung Choi, Hee-Jo Koo, Hyeri Seok, Ji Hoon Jeon, Won Suk Choi, Dae Jung Kim, Dae Won Park, Euna Han

## Abstract

**Background:** Angiotensin receptor blockers (ARBs) and angiotensin converting enzyme inhibitors (ACEIs) have anti-inflammatory effects. Severe acute respiratory syndrome coronavirus 2 (SARS-CoV-2) uses the membrane protein angiotensin-converting enzyme 2 (ACE2), which is increased by ARB/ACEI treatment, as a cell entry receptor. Therefore, the use of ARBs/ACEIs for COVID-19 remains controversial.

**Methods:** A retrospective case-control study was conducted using COVID-19 patients previously diagnosed with hypertension before COVID-19 onset. The primary outcome was severe infection or all-cause mortality. Cases included ARB/ACEI use for ≥30 days during the 6 months before COVID-19 onset. Primary controls included antihypertensive use other than ARBs/ACEIs (narrow control); secondary controls included all other hypertension patients (broad control). We investigated ARB/ACEI association with outcomes in general and by subgroups (age, sex, and presence of diabetes) using logistic regression models with propensity score matching.

**Findings:** Of 234427 suspected COVID-19 patients we screened, 1585 hypertension patients were analyzed. In the 892 cases, 428 narrow controls, and 693 broad controls, severe infection or death occurred in 8·6%, 22·2%, and 16·7%, respectively. ARB/ACEI use was associated with a reduced risk of severe infection or death relative to the narrow control group (adjusted odds ratio [aOR] 0·43, 95% confidence interval [CI] 0·28 – 0·65) and broad control group (aOR 0·49, 95% CI 0·33 – 0·71). The association was smaller for newly diagnosed hypertension patients (aOR 0·11, 95% CI 0·03 – 0·42 compared to narrow control group). ARB/ACEI protective effects against severe infection or death were significantly observed in male and diabetic patients.

**Interpretation:** ARB/ACEI use was associated with a lower risk of severe infection or mortality compared to other antihypertensives or ARB/ACEI nonuse.

**Funding:** None

**Research in context:** *Evidence before this study:* Animal studies reported that ACE2 attenuates lung injury and provides a protective effect against severe pneumonia. Additionally, retrospective studies found that ARBs/ACEIs may have beneficial effects on ARDS patient survival. Previous observational studies have reported no potential harmful association of either ARBs or ACEIs with COVID-19 outcomes.

*Added value of this study:* By analyzing nationwide claims data in South Korea, we found that previous use of ARB/ACEI was associated with improved outcomes in COVID-19 compared with either nonuse or use of a different class of antihypertensive drugs. The risk of severe infection or death was consistently about 55% lower in those treated with ARB/ACEIs than those who were not exposed to ARB/ACEIs. The protective effect of ARB/ACEI was remained significantly among the male subgroup and patients with diabetes. This association was also observed among COVID-19 patients with newly diagnosed hypertension.

*Implications of all the available evidence:* These results provide supporting evidence for the continued use of ARBs/ACEIs among patients with COVID-19. Moreover, for newly diagnosed hypertension patients, initiation of ARB/ACEI use may not adversely affect COVID-19 prognosis. Given the poor prognosis of COVID-19 patients with hypertension and lack of curable strategy, these findings may have considerable clinical implications in prevention of poor outcome in patients with hypertension.

## Introduction

Severe acute respiratory syndrome coronavirus 2 (SARS-CoV-2), which causes coronavirus disease 2019 (COVID-19), is similar to SARS-CoV in 2003.^1^ COVID-19 patients are at risk of acute respiratory distress syndrome (ARDS) and death.^2^ Although vaccines and drugs are being developed, investigations to enhance the supportive care of high-risk patients is also warranted. Recent studies of COVID-19 patients in China have reported more severe disease outcomes in patients with hypertension.^2-4^ Angiotensin receptor blockers (ARB) and angiotensin-converting enzyme inhibitors (ACEI) are recommended primary agents for hypertension management.^5^ Angiotensin-converting enzyme 2 (ACE2) is an enzyme that physiologically counters the activation of the renin-angiotensin-aldosterone system (RAAS) and a recently identified co-receptor for SARS-CoV-2 viral entry.^6^ Some studies suggest that RAAS inhibitors, such as ARBs and ACEIs, increase ACE2 expression.^7^ Therefore, concerns have been raised about the effects of RAAS inhibitors in severe COVID-19 infection.^8,9^ Consequently, patients and physicians in the highly epidemic region of COVID-19 have become concerned about the continued use or the initiation of RAAS inhibitor for previously prescribed or newly diagnosed hypertension patients.

Conversely, other medical professionals hypothesize that use of ARBs or ACEIs may reduce COVID-19 severity.^10,11^ This expectation is based on evidence of host response effects instead of antiviral effects. ARBs and ACEIs reduced cytokine levels in lung injury models^12,13^ and are associated with improved pneumonia outcomes.^14^ While there is a lack of evidence suggesting that ARBs or ACEIs decrease SARS-CoV-2 induced lung injury, these previous studies suggest that ARBs or ACEIs have potential protective effects in COVID-19. Due to the lack of clear evidence for the effects of ARBs or ACEIs on COVID-19 prognosis, the Council on Hypertension of the European Society of Cardiology and the American Heart Association recommended not to change antihypertensive drug use solely for COVID-19.^1^Some observational studies also reported that ARBs/ACEIs did not increase the likelihood of a positive COVID-19 test^16,17^ or promote the progression of severe COVID-19.^16-20^ However, these results cannot be generalized due to differences in national response systems and COVID-19 situations. Furthermore, most of the previous studies were not based on the nationwide data.

Here, to investigate the association between ARB or ACEI (ARB/ACEI) use and COVID-19 severity, we conducted a retrospective case-control study using nationwide insurance claims data in South Korea, wherein COVID-19 incidence rate was 21·25 per 100000 persons as of midnight on May 15, 2020.^21^

## Methods

### Data source

We used insurance claim data from the Health Insurance Review and Assessment Service (HIRA) database for National Health Insurance in South Korea, under which all legal residents are compulsory beneficiaries and all medical institutions and pharmacies are mandatory providers. The HIRA database contains all information regarding the diagnosis, prescribed medications, and medical procedures of approximately 50 million Koreans. The Ministry of Health and Welfare of Korea and HIRA initiated the #OpenData4Covid19 project, a global research collaboration on COVID-19.^22^ The data set is based on insurance claims sent to HIRA by May 15, 2020, and comprised of a complete three-year medical service history of all COVID-19 tested cases.

### Study outcomes

For this study, we focused on the primary composite endpoint of severe infection or death. Similar to previous studies that assessed the COVID-19 severity,^3,23,24^ patients with one of the followings were considered as a severe infection case: 1) respiratory failure requiring mechanical ventilation, 2) organ failure requiring admission to the intensive care unit (ICU), 3) organ failure requiring continuous renal-replacement therapy (CRRT), or 4) organ failure requiring extracorporeal membrane oxygenation (ECMO) treatment. Associated procedure codes are listed in Table S1 in the Supplementary Appendix. Secondary endpoints were all-cause mortality and severe infection, respectively.

### Case definitions

Confirmed COVID-19 patients was identified by variable generated by linkage from Korea Centers for Disease Control and Prevention (KCDC).

Among identified COVID-19 patients, patients with concomitant hypertension were defined as those with hypertension diagnostic codes (ICD-10: I10, I11, I12, I13, or I15) at least once during six months or those prescribed antihypertensive drugs (the Anatomical Therapeutic Chemical (ATC) classification code: C02)^25^ for more than 30 consecutive days prior to COVID-19 onset. The study sample was divided into ARB/ACEI group and control groups according to their antihypertensive medications before COVID-19 onset. Specifically, hypertension patients who used ARBs or ACEIs for ≥ 30 days during the 6 months prior to COVID-19 onset were defined as the case group. Hypertension patients prescribed alternative antihypertensive drugs for at least 30 days within 6 months prior to COVID-19 onset were defined as the primary control group (narrow group, hereafter). As the second control group, hypertension patients not prescribed antihypertensive drugs, or prescribed antihypertensive drugs for less than 30 days, were also added to the narrow control group (broad control group, hereafter). Patients with antihypertension drug prescription changes e.g., from case to narrow control, or from narrow control to case group) during COVID-19 treatment were excluded from analysis. (N = 70). However, patients who ceased antihypertensive medication use without changing its type during COVID-19 treatment remained in the analysis. (Figure 1)

**Figure 1.**
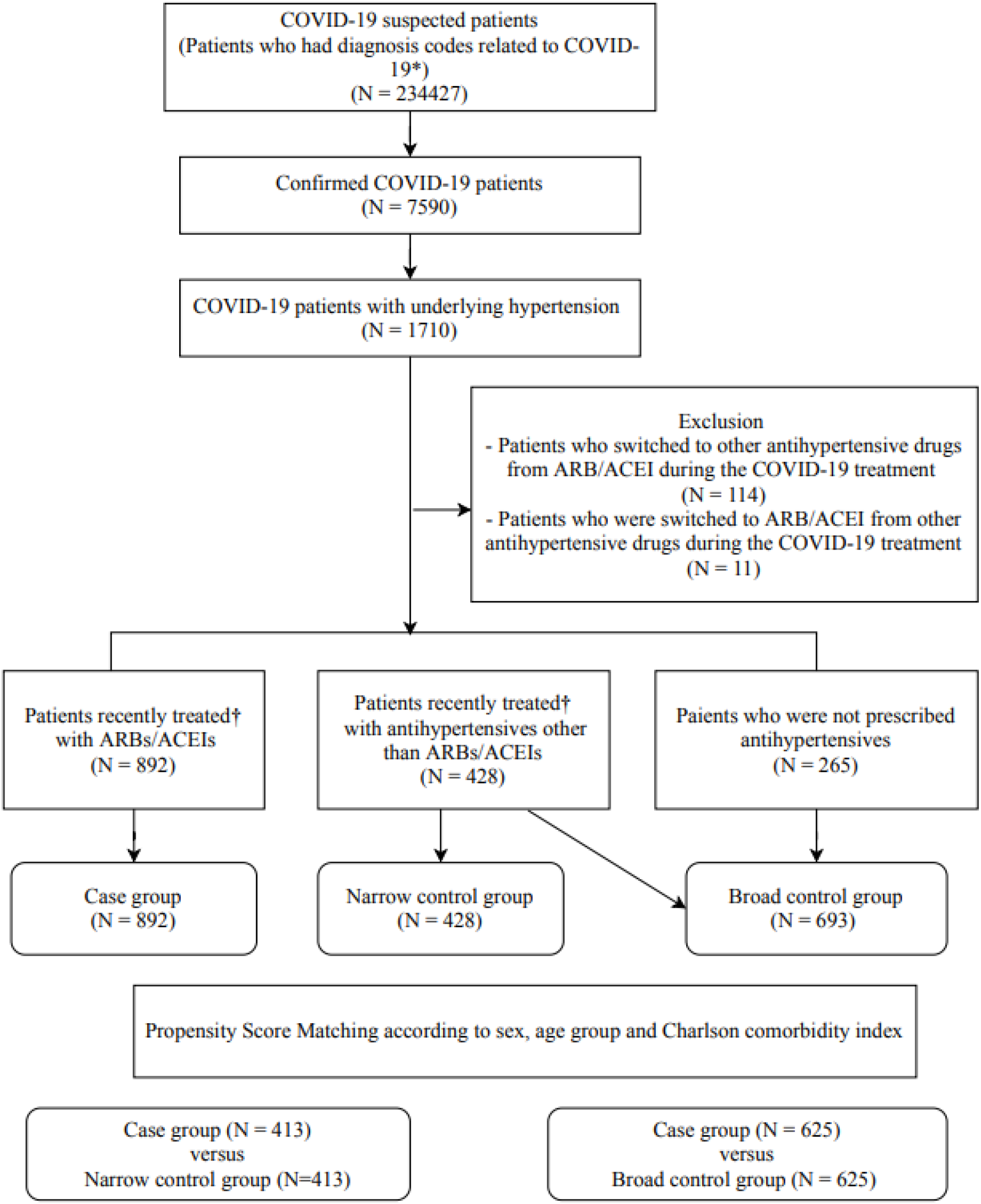
Flow Diagram of the Study Population. * Diagnosis codes related to COVID-19 are B34.2 (Coronavirus infection, unspecified), B97.2 (Coronavirus as the cause of disease classified to other chapters), U18 (Provisional assignment of new diseases or emergency use at a national level), U18.1 (Novel coronavirus infection), or U07.1 (COVID-19). † Recent antihypertensive drug treatment was defined as prescribed for at least 30 days during the 6 months prior to COVID-19 onset. ARB denotes Angiotensin receptor blocker and ACEI angiotensin-converting enzyme inhibitor.

Among the confirmed COVID-19 patients, we also identified patients with hypertension newly diagnosed in the six months prior COVID-19 onset. Newly diagnosed hypertension patients were defined as subjects without any previous claims associated with hypertension, including antihypertensive medications or diagnosis code, during the past 6 months.

### Covariates

Demographic information (sex and age) were controlled as covariates. We also extracted regional information of the medical facilities that treated the COVID-19 patients. We controlled for the presence of selected comorbidities during the 3 years prior to COVID-19 onset and during COVID-19 treatment. Comorbidities were defined based on ICD-10 codes: diabetes mellitus (ICD-10 codes: E10–E14), chronic lung disease [asthma (J45), chronic obstructive pulmonary disease (J44), and emphysema (J43)], and major neurologic disease^26^ [cerebrovascular disease (I60-I69), epilepsy (G40-G41), Parkinson’s disease (G20-G22), and dementia (G30-G31, F00-F03)]. Use of Lopinavir/ritonavir (ATC code: J05AR10) or hydroxychloroquine (ATC code: P01BA02) during COVID-19 treatment was also controlled for as a covariate, as these drugs were recommended first-line therapies for COVID-19 in South Korea.^27^ In addition, the Charlson comorbidity index was constructed and controlled for in all estimations (associated ICD-10 codes are listed in Table S2 in the Supplementary Appendix).

### Subgroup analyses

As the ACE2 expression is more likely reduced in older patients,^28-30^ males,^29,30^ and diabetic patients,^30^ subgroup analyses were conducted by sex (male versus female), age (<65 years versus ≥65 years), and diabetes.

### Statistical analysis

Continuous variables were expressed as mean (standard deviations) and the ARB/ACEI group was compared to the control group using t-tests. Categorical variables were summarized as counts (percentages) and distributional differences were analyzed by chi-squared tests or Fisher’s exact tests comparing the ARB/ACEI group versus the control group. A multivariate logistic regression analysis was performed to investigate the impact of ARB/ACEI use on primary and secondary outcomes. The multivariate models were adjusted for the aforementioned covariates.

We observed difference in baseline clinical characteristics among case and control groups and developed propensity score matched cohorts to control potential confounders associated with the exposure to ARB/ACEI, including age group (10-year interval), sex, and Charlson comorbidity index. For propensity score matching, a greedy nearest neighbor matching scheme was used. The balance of covariates was evaluated by estimating standardized differences (SD) before and after matching, and small absolute value less than 0.1 was considered successful balancing between the two groups. ARB/ACEI and control groups were paired at 1:1, respectively.

All analyses were performed with SAS software, version 9·4 (SAS Institute, Cary, NC, USA), and a 2-tailed P < 0·05 was used to indicate statistical significance. The 95% CI has not been adjusted for multiple testing and should not be used to infer definitive effects.

The present study was approved by Yonsei institutional review boards (7001988-202004-HR-843-01E.)

## Results

### Patients and treatment of COVID-19

All of the 234427 suspected COVID-19 patients had diagnosis codes related to COVID-19. Of these, 7590 patients were classified as confirmed COVID-19 patients. A total of 1710 (22·5%) of the confirmed COVID-19 patients had underlying hypertension. After exclusion, our final study sample was composed of 1585 patients with hypertension and COVID-19 infection. The patients were subdivided into the following three groups: ARB/ACEI group (892 patients), narrow control group (428 patients), and broad control group (693 patients). After propensity score matching, 413 and 625 of 892 patients in the ARB/ACEI group were successfully matched to narrow control group and broad control group, respectively (Figure 1).

Table 1 presents the baseline characteristics and treatment distributions for the entire sample and by hypertension treatment groups. The ARB/ACEI group was statistically significantly younger, and had a lower proportion of major neurologic diseases, than the control groups. Charlson comorbidity index was lower in the ARB/ACEI group than the control groups. Of all COVID-19 sample patients, approximately 20% had chronic lung diseases, and about half were diagnosed with diabetes. These comorbidities were not statistically significantly different across groups. During COVID-19 associated treatments, about two-thirds of patients were admitted in hospitals located in a metropolitan area. However, no statistically significant differences concerning location were observed between the ARB/ACEI group and the control groups. Lopinavir/ritonavir was administered in 840 (53·0%) patients and hydroxychloroquine was administered in 737 (46·5%) patients among the COVID-19 patients with hypertension. Hydroxychloroquine was more frequently administered to control groups compared to ARB/ACEI group.

**Table 1:** Demographics and clinical characteristics of COVID-19 patients with hypertension.

| Characteristic | ARB/ACEI<br>(N = 892) | Narrow control<br>(N = 428) | Broad control<br>(N = 693) | p<br>value† | p<br>value‡ |
| --- | --- | --- | --- | --- | --- |
| Age – yr | 65 ± 13 | 71 ± 13 | 68 ± 15 | < 0.001 | < 0.001 |
| Age group – no. (%) |  |  |  | < 0.001 | < 0.001 |
| Age <65 years | 464 (52.0) | 129 (30.1) | 278 (40.1) |  |  |
| Age ≥ 65 years | 428 (48.0) | 299 (69.9) | 415 (59.9) |  |  |
| Sex – no. (%) |  |  |  | 0.58 | 0.91 |
| Male | 381 (42.7) | 176 (41.1) | 298 (43.0) |  |  |
| Female | 511 (57.3) | 252 (58.9) | 395 (57.0) |  |  |
| Region of hospitals – no. (%) |  |  |  | 0.46 | 0.80 |
| Metropolitan | 506 (56.7) | 258 (60.3) | 388 (56.0) |  |  |
| Urban | 348 (39.0) | 152 (35.5) | 279 (40.3) |  |  |
| Rural | 38 (4.3) | 18 (4.2) | 26 (3.8) |  |  |
| Comorbidities – no. (%) |  |  |  |  |  |
| Charlson comorbidity index | 0.87 ± 1.19 | 1.00 ± 1.16 | 0.94 ± 1.16 | 0.06 | 0.25 |
| Diabetes | 415 (46.5) | 178 (41.6) | 297 (42.9) | 0.09 | 0.15 |
| Major neurologic diseases | 250 (28.0) | 202 (47.2) | 296 (42.7) | < 0.001 | < 0.001 |
| Chronic lung diseases | 173 (19.4) | 87 (20.3) | 136 (19.6) | 0.69 | 0.91 |
| Treatment – no. (%) |  |  |  |  |  |
| Lopinavir/ritonavir | 473 (53.0) | 247 (57.7) | 367 (53.0) | 0.11 | 0.98 |
| Hydroxychloroquine | 384 (43.1) | 235 (54.9) | 353 (50.9) | < 0.001 | 0.001 |
| ICU care | 33 (3.7) | 44 (10.8) | 53 (7.7) | < 0.001 | < 0.001 |
| Ventilator care | 22 (2.5) | 28 (6.5) | 29 (4.2) | < 0.001 | 0.05 |
| ECMO | 5 (0.6) | 1 (0.2) | 1 (0.1) | 0.41 | 0.18 |
| CRRT | 3 (0.3) | 8 (1.9) | 8 (1.2) | 0.004 | 0.05 |

ARB denotes Angiotensin receptor blocker, ACEI angiotensin-converting enzyme inhibitor, ICU intensive care
unit, ECMO extracorporeal membrane oxygenation, CRRT continuous renal-replacement therapy, OR odds ratio, and CI confidence interval.
† Denotes P value between ARB/ACEI and narrow control as quantified by t-test
‡ Denotes P value between ARB/ACEI and broad control as quantified by t-test

### Primary of outcome

Among COVID-19 patients with concomitant hypertension, the primary composite endpoint event occurred in 193 patients (12·2 %); 77 (8·6%) in the ARB/ACEI group, 95 (22·2%) in the narrow control group, and 116 (16·7%) in the broad control group. The risk of severe infection or death was significantly lower in the ARB/ACEI group versus the narrow control group (*P* < 0·001) or the broad control group (*P* < 0·001). After adjusted by a multivariate logistic regression model, patients prescribed with ARBs/ACEIs had a reduced risk of severe infection or death relative to the narrow control group (adjusted odds ratio [aOR] 0·43, 95% confidence interval [CI] 0.30 – 0.75) and broad control group (aOR 0.54, 95% CI 0.38 – 0.75). The primary outcome of the matched population showed significantly lower rate in the ARB/ACEI group compared to narrow control group (aOR 0·43, 95 % CI 0·28 – 0·65) (Table 2) and broad control group (aOR 0·49, 95% CI 0·33 – 0·71) (Table 3).

**Table 2:** Study outcomes in propensity score-matched ARB/ACEI versus narrow control group.

|  | **ARB/ACEI group**<br>(N = 413) | **Narrow control group**<br>(N = 413) | **P value** | **aOR (95% CI)\*** |
| --- | --- | --- | --- | --- |
| Severe infection or death | 53 (12.8) | 93 (22.5) | < 0.001 | 0.43 (0.28 – 0.65) |
| Severe infection | 25 (6.1) | 53 (12.8) | < 0.001 | 0.37 (0.22 – 0.63) |
| Death | 39 (9.4) | 65 (15.7) | 0.006 | 0.53 (0.31 – 0.90) |

\*Adjusted for age, sex, region of hospitals, comorbidities (diabetes, chronic lung disease, and major neurologic diseases), Charlson comorbidity index, and treatment modalities. ARB denotes Angiotensin receptor blocker, ACEI angiotensin-converting enzyme inhibitor, aOR adjusted odds ratio, and CI confidence interval.

**Table 3:** Study outcomes in propensity score-matched ARB/ACEI versus broad control group.

|  | **ARB/ACEI group**<br>(N = 625) | **Broad control group**<br>(N = 625) | **P value** | **aOR (95% CI)\*** |
| --- | --- | --- | --- | --- |
| Severe infection or death | 63 (10.1) | 102 (16.3) | 0.001 | 0.49 (0.33 – 0.71) |
| Severe infection | 34 (5.4) | 55 (8.8) | 0.02 | 0.49 (0.30 – 0.79) |
| Death | 42 (6.7) | 69 (11.0) | 0.007 | 0.48 (0.29 – 0.79) |

Among the patients with newly diagnosed hypertension, the ARB/ACEI group was associated with a lower risk of severe infection or death compared to the narrow control group (aOR 0.16, 95% CI 0.06 – 0.43) and the broad control group (aOR 0.28, 95% CI 0.11 – 0.67) (Table S3 in the Supplementary Appendix). After propensity score matching, the protective effect of the ARB/ACEI among patients with newly diagnosed hypertension remained statistically significant with an adjusted odds ratio of 0·11 (95% CI 0·03 – 0·42) compared to narrow control group. Comparison with the broad control group was not performed because propensity score matching was not feasible.

### Secondary outcomes

In COVID-19 patients diagnosed with hypertension, the severe infection rate in the ARB/ACEI group was lower than the narrow control group (4·9% vs. 12·6%, *P* < 0·001) and broad control group (4·9% vs. 9.1%, *P* = 0·001). In the multivariate logistic regression, ARB/ACEI use was associated with a lower risk of severe infection relative to the narrow control group (aOR 0·40, 95% CI 0·25 – 0·62) and broad control group (aOR 0·51, 95% CI 0·33 – 0·78). The lower risk of severe infection remained consistent and statistically significant in the propensity score-matched analysis (aOR 0·37, 95% CI 0·22 – 0·63 compared to narrow control group, Table 2; aOR 0·49, 95% CI 0·30 – 0·79 compared to broad control group, Table 3).

In the ARB/ACEI group, the all-cause mortality rate was lower than in the narrow control group (5·5% vs. 15·7%, *P* < 0·001) and broad control group (5·5% vs. 11·4%, *P* < 0·001). The lower risk of all-cause mortality in the ARB/ACEI group remained consistent in the adjusted analyses using multivariate logistic regression model (aOR 0·48, 95% CI 0·30 – 0·77 compared to the narrow control; aOR 0·60, 95% CI 0·39 – 0·95 compared to the broad control group) and propensity score matched analysis (aOR 0·53,95% CI 0·31 – 0·90 compared to the narrow control, Table 2; aOR 0·48, 95% CI 0·29 – 0·79 compared to the broad control group, Table 3).

Among newly diagnosed hypertension patients, ARB/ACEI use was consistently associated with a lower risk of severe infection (aOR 0·12, 95% CI 0·04– 0·38 compared to the narrow control; aOR 0·21, 95% CI 0·07 – 0·62 compared to the broad control group; aOR 0·04, 95% CI 0·01– 0·35 compared to the narrow control with propensity score matching) (Table S3 in the Supplementary Appendix). However, the all-cause mortality of the ARB/ACEI group was not significantly different than the control groups (Table S3 in the Supplementary Appendix).

### Subgroup analysis

Subgroup analysis was performed for COVID-19 patients with hypertension with propensity score matched sets. In the male subgroup, the use of ARBs/ACEIs was correlated with a reduced risk of severe infection or death, when compared to the narrow control group (aOR 0·26, 95% CI 0·13 – 0·49) and broad control group (aOR 0·38, 95% CI 0·22 – 0·66). However, this correlation did not reach statistical significance in the female subgroup (aOR 0·69, 95% CI 0·69 – 1·21 compared to the narrow control group; aOR 0·62, 95% CI 0·36 – 1·06 compared to the broad control group) (Table 4). Similarly, the reduced odds of ARB/ACEI group for secondary outcomes was observed only in the male subgroup.

**Table 4:**
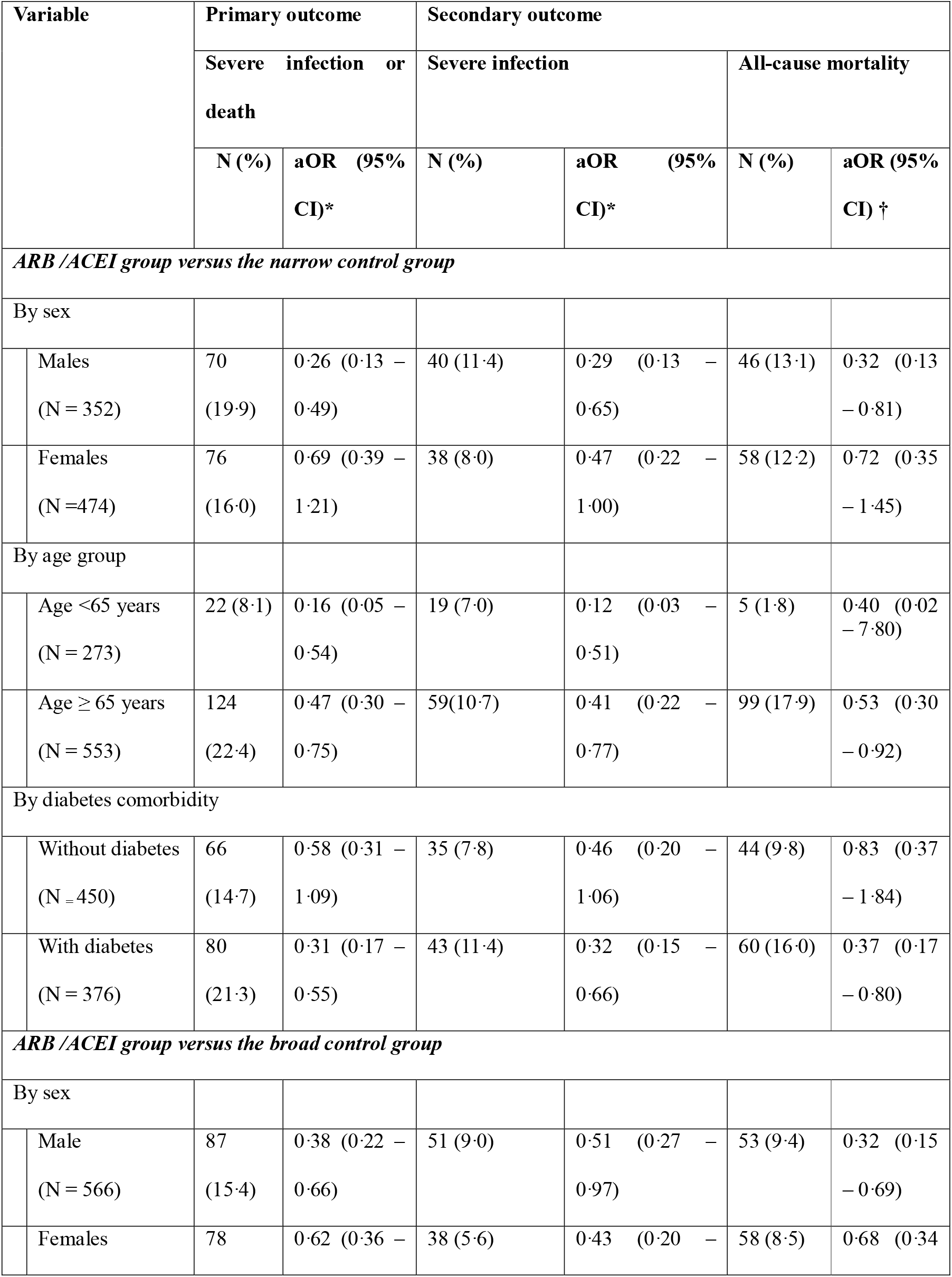
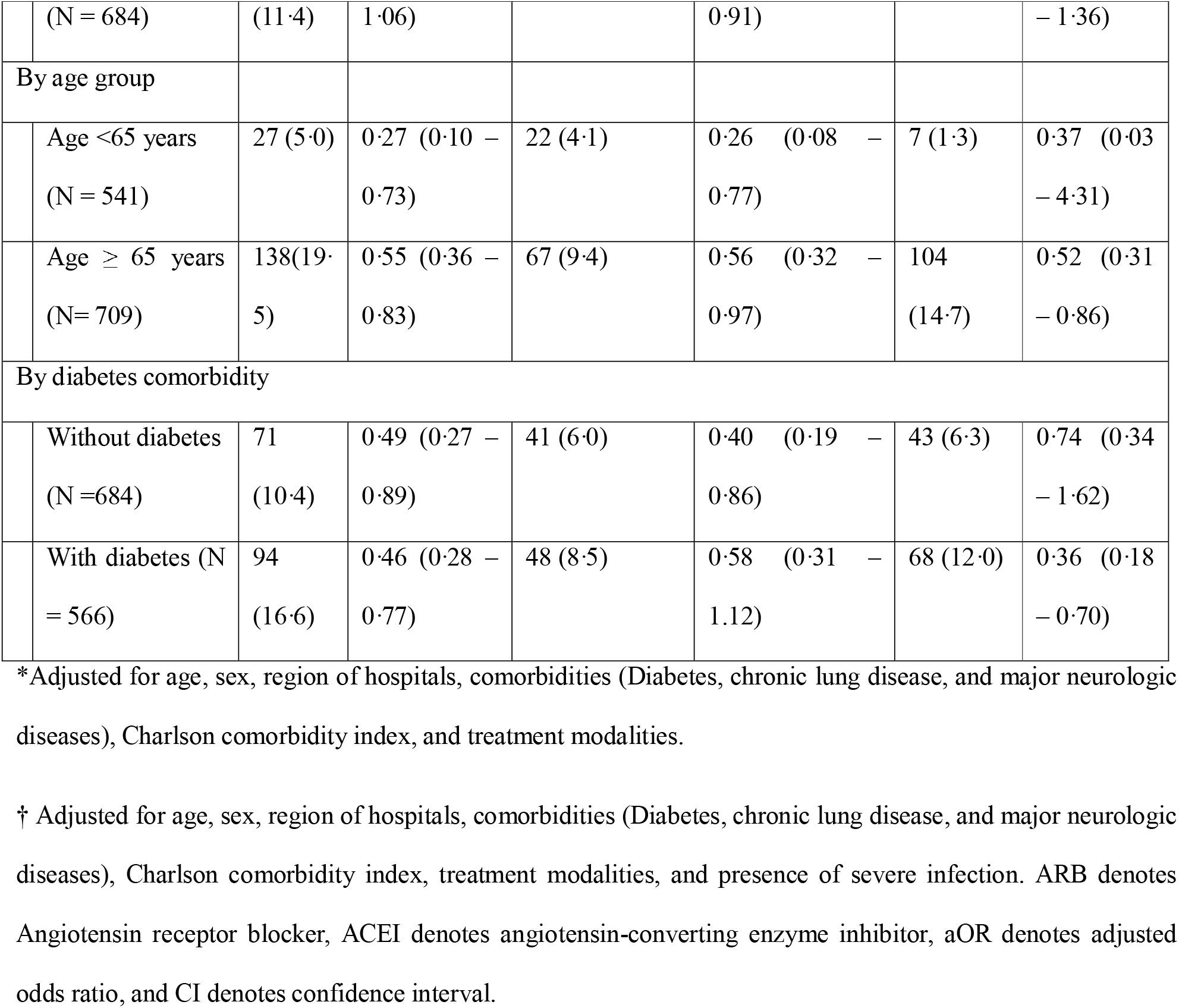
Subgroup analysis according to sex, age group, and diabetes comorbidity with propensity score matched set.

Both the younger subgroup and older age subgroup analysis showed that the ARB/ACEI group had a statistically significantly reduced odds ratio for primary outcome and severe infection, compared to unexposed groups (Table 4).

Among the non-diabetic subgroup, the use of ARBs/ACEIs showed a reduced risk of severe infection or death compared to broad control group (aOR 0·49, 95% CI 0·27 – 0·89). However, the association was not statistically significant when compared to narrow control group (aOR 0·58, 95% CI 0·31 – 1·09). In the subgroup with diabetes, ARB/ACEI use was consistently and significantly associated with a lower risk of severe infection or death (aOR 0·31, 95% CI 0·17 – 0·55 compared to the narrow control group; aOR 0·46, 95% CI 0·28 – 0·77 compared to the broad control group) (Table 4). The ARB/ACEI group in the diabetic patients also showed a reduced risk for secondary outcomes compared to other groups.

We also analyzed the subgroup sets without propensity score matching and the results are shown in the Table S4 in the Supplementary Appendix.

## Discussion

This study shows that ARB/ACEI use is associated with improved outcomes in COVID-19 patients compared with either nonuse or use of a different class of antihypertensive drugs among patients with hypertension. This association was also observed among COVID-19 patients with newly diagnosed hypertension. These results provide supporting evidence for the continued use of ARBs/ACEIs among patients with COVID-19. Moreover, for newly diagnosed hypertension patients, initiation of ARB/ACEI use may not adversely affect COVID-19 prognosis.

ARBs and ACEIs up-regulate ACE2. In an experimental animal study using rats, intravenous infusions of ARBs and ACEIs increased ACE2 receptor expression in cardiopulmonary circulation.^31^ Therefore, patients taking ARBs/ACEIs are presumed to have elevated ACE2 receptor levels in cardiopulmonary circulation.^8^ Importantly, the tissue binding sites for the anchoring spike proteins of the COVID-19 is ACE2.^1^ Consequently, patients treated with ARBs/ACEIs may be at increased risk of severe COVID-19.^8,9^

Meanwhile, many have hypothesized that elevated ACE2 expression levels may be beneficial rather than harmful in patients with COVID-19.^10^ Animal studies reported that ACE2 attenuates lung injury and provides a protective effect against severe pneumonia.^12,13^ An observational study that examined patients with respiratory syncytial virus infection reported that higher ACE2 levels are associated with reduced severity of ARDS.^32^ Additionally, a retrospective case-control study found that ARBs/ACEIs may have beneficial effects on ARDS patient survival.^33^

Previous observational studies^16-20,34^ have reported no potential harmful association of either ARBs or ACEIs with COVID-19 outcomes. In the current study, recent ARB/ACEI use did not increase the risk of severe infection or death. More importantly, we observed potentially beneficial protective effects in COVID-19 patients previously prescribed ARBs/ACEIs. The risk of severe infection or death was consistently about 55% lower in those treated with ARB/ACEIs than those who were not exposed to ARB/ACEIs. This association persisted even with propensity score matching, multiple subgroup analysis and adjustment for available confounders. We could not fully explain why ARBs/ACEIs use showed better outcomes in the present study. As ARBs/ACEIs are not antiviral agents for the SARS-CoV-2, the effects of these drugs may be due to factors other than the COVID-19, such as hypertension control itself. Although we included patients who regularly visited clinics with hypertension more than once during the 6 months before the COVID-19 onset, broad control group may have both mild hypertensive patients and poor adherence patients. Comparing the ARB/ACE to narrow control group with other antihypertensive prescribed would mitigate such confounding effects, but responses to treatment, such as side-effects or contraindications would affect whether patients be treated with which hypertension treatment option. That is, assignment to each treatment option for hypertension was not random in the present study. Randomized controlled trials would ideally overcome these shortcomings despite the ethical issue of randomization.

A priori, we hypothesized that as the ACE2 expression is more likely to decrease in older age,^28-30^ male sex^29,30^ and diabetic patients,^30^ ARB/ACEI use would affect COVID-19 outcomes differently according to these subgroups. As such, we found that ARB/ACEI-mediated protective effects were more prominent in the males and diabetic patients, which are known to have less ACE2 expression than the other groups.

We obtained all potential COVID-19 cases in Korea from 20 January 2020 until 15 May 2020, using national insurance claims with all Koreans as compulsory beneficiaries. Moreover, we could assess the confirmed COVID-19 cases by using updated data. HIRA data was not linked to KCDC data and only claims of suspected COVID-19 patients were provided, without information whether the COVID-19 was confirmed. However, from 27 May 2020, HIRA data was linked to KCDC’s confirmed case data up to 15 May 2020 and we analyzed this updated data. Therefore, the findings of the present study may be extrapolated as evidence for the whole Korean population. An important aspect of this study is that individual patients can be followed up until death unless they lose quantification of the National Health Insurance of Korea due to emigration, for example. Hence, we accurately collected all past medical history and prognosis in the present study, and could control for confounding variables such as Charlson comorbidity index as a covariate.

This study has some limitations due to the characteristics of the claims data. First, a patient’s socioeconomic status-related information was not included. However, as the entire cost of COVID-19 treatment is paid by the NHIS, we do not expect any substantial variations due to socioeconomics in regards to the impact of ARBs/ACEIs on the COVID-19 treatment. Second, data on participant smoking behavior, which is regarded as a major risk factor for severe COVID-19,^35,36^ was unavailable for analysis. Third, the effects of the degree and duration of ARB/ACEI use on COVID-19 outcomes were not fully assessed. Although a protective effect was observed in even newly diagnosed hypertension patients, more well-designed and controlled studies investigating the protective capabilities of ARB/ACEI treatment on COVID-19 prognosis are needed.

In conclusion, the recent use of ARBs or ACEIs was significantly correlated with reduced risk of poor COVID-19 outcomes relative to either use of a different class of antihypertensive medication or nonuse of ACEIs/ARBs among patients with hypertension.

## Data Availability

The Ministry of Health and Welfare of Korea and HIRA initiated the #OpenData4Covid19 project, a global research collaboration on COVID-19 via https://covid19data.hira.or.kr. Researchers can directly request data from this site.

## Acknowledgments

The authors appreciate healthcare professionals dedicated to treating COVID-19 patients in Korea, and the Ministry of Health and Welfare and the Health Insurance Review & Assessment Service of Korea for sharing invaluable national health insurance claims data in a prompt manner.

## Supplementary Appendix

**Supplementary Table S1.** Procedure codes associated with definitions.

| Procedure | Associated codes |
| --- | --- |
| Intensive care unit | AJ001, AJ003, AJ004, AJ05, AJ006, AJ007, AJ008, AJ009, AJ010, AJ011, AJ020, AJ021, AJ031, AJ041, AJ042, AJ043, AJ044, AJ045, AJ046, AJ100, AJ101, AJ102, AJ103, AJ110, AJ111, AJ112, AJ120, AJ121, AJ122, AJ130, AJ131, AJ132, AJ140, AJ141, AJ142, AJ143, AJ144, AJ150, AJ151, AJ152, AJ160, AJ161, AJ180, AJ190, AJ200, AJ201, AJ203, AJ210, AJ211, AJ212, AJ220, AJ221, AJ22, AJ230, AJ231, AJ232, AJ240, AJ241, AJ242, AJ244, AJ250, AJ251, AJ252, AJ260, AJ261, AJ261, AJ280, AJ290, AJ300, AJ301, AJ301, AJ302, AJ303, AJ310, AJ311, AJ312, AJ312, AJ320, AJ321, AJ330, AJ331, AJ332, AJ340, AJ341, AJ342, AJ350, AJ351, AJ352, AJ360, AJ380, AJ390, AJ500, AJ510, AJ520, AJ530, AJ540, AJ550, AJ560, AJ580, AJ590 |
| Mechanical ventilation | M0850, M0857, M0858, M0860, M5830, M5850, M5851, M5852, M5853, M5854, M5855, M5856, M5857, M5858, M5860 |
| Continuous renal-replacement therapy | O7031, O7031, O7033, O7034 |
| Extracorporeal membrane oxygenation | O1901, O1902, O1903, O1904 |

**Supplementary Table S2.** Charlson comorbidity index and associated ICD-10 codes.

| Variables | ICD-10 codes |
| --- | --- |
| AIDS/HIV | B20-B24 |
| Cerebrovascular disease | I60-I69,G45-46 |
| Congestive heart failure | I09.9, I11.0, I13.0, I13.2, I25.5, I42-43, I50 |
| Chronic pulmonary disease | I27.8, I27.9, J68.4, J70.1, J70.3, J40-J47, J60-J67 |
| Dementia | F00-F03, F05.1, G30, G31.1 |
| Diabetes without chronic complication | E10.0, E10.1, E10.6, E10.8, E10.9, E11.0, E11.1 E11.6, E11.8, E11.9, E12.0, E12.1, E12.6, E12.8, E12.9, E13.0, E13.1,E13.6, E13.9, E14.0, E14.1, E14.6, E14.8, E14.9 |
| Diabetes with chronic complication | E10.2-E10.5, E11.2-E11.5, E11.7, E12.2-E12.5, E12.7, E13.2-E13.5, E13.7, E14.2-E14.5, E14.7 |
| Hemiplegia or paraplegia | G81-G82,<br>G04.1,G11.4,G80.1,G80.2,G83.0,G83.1,G83.2,G83.3,G83.4,G83.9 |
| Mild liver disease | B18, K70.0, K70.1, K70.2, K70.3, K70.9, K71.3, K71.4, K71.5, K71.7, K73-K74, K76.0, K76.2,K76.3, K76.4,K76.8, K76.9, Z94.4 |
| Moderate or severe liver disease | K72.1, K72.9, K76.6, K76.7 |
| Any malignancy, including leukemia and lymphoma | C00-C26, C30 -C34, C37-C41, C43, C45-C58, C60-C76., C81-C85, C88, C90-C96 |
| Metastatic solid tumor | C77-C80 |
| Myocardial infarction | I25.2, I21-I22 |
| Peripheral vascular disease | I70-I71, I73.1, I73.9, I77.1, I79.0, I79.2, K55.1,K55.8, K55.9, Z95.8, Z95.9 |
| Peptic ulcer disease | K25- K28 |
| Rheumatologic disease | M05- M06, M32- M34, M31.5, M35.1, M35.3, M36.0 |
| Renal disease | I12.0, I13.0, N18-N19, N25.0, Z94.0,Z99.2,N03.3,N03.4,N03.5,N03.6,N03.7,N05.2, N053,N05.4,N05.5,N05.6,N05.7,Z49.0,Z49.1,Z49.2 |

**Supplementary Table S3.**
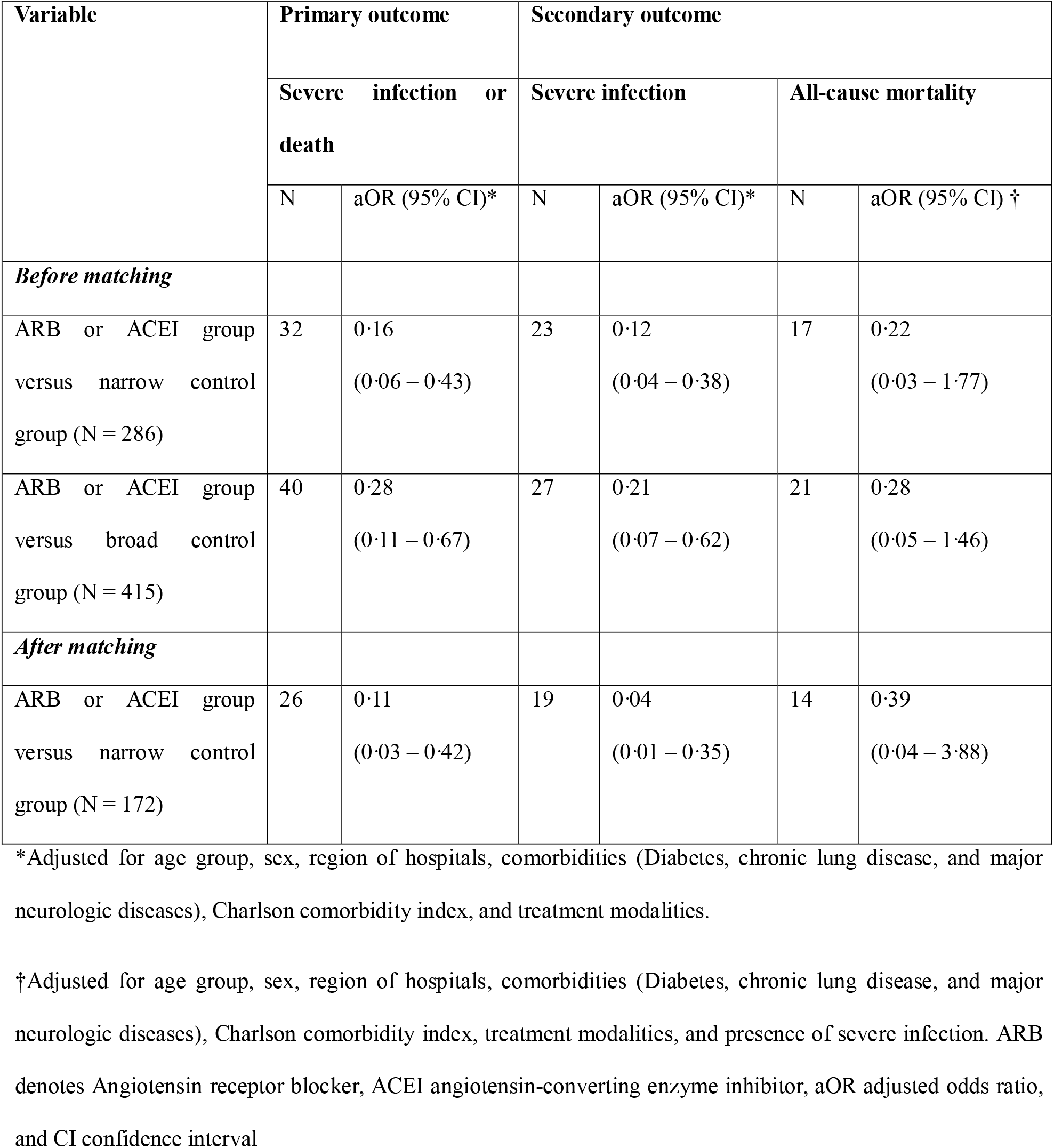
Association between antihypertensive medication use and clinical outcomes in COVID-19 patients with newly diagnosed hypertension.

| Variable | Primary outcome |  | Secondary outcome |  |  |  |
| --- | --- | --- | --- | --- | --- | --- |
|  | Severe infection or death |  | Severe infection |  | All-cause mortality |  |
|  | N | aOR (95% CI)* | N | aOR (95% CI)* | N | aOR (95% CI) † |
| ***Before matching*** |  |  |  |  |  |  |
| ARB or ACEI group versus narrow control group (N = 286) | 32 | 0.16<br>(0.06 – 0.43) | 23 | 0.12<br>(0.04 – 0.38) | 17 | 0.22<br>(0.03 – 1.77) |
| ARB or ACEI group versus broad control group (N = 415) | 40 | 0.28<br>(0.11 – 0.67) | 27 | 0.21<br>(0.07 – 0.62) | 21 | 0.28<br>(0.05 – 1.46) |
| ***After matching*** |  |  |  |  |  |  |
| ARB or ACEI group versus narrow control group (N = 172) | 26 | 0.11<br>(0.03 – 0.42) | 19 | 0.04<br>(0.01 – 0.35) | 14 | 0.39<br>(0.04 – 3.88) |

\*Adjusted for age group, sex, region of hospitals, comorbidities (Diabetes, chronic lung disease, and major neurologic diseases), Charlson comorbidity index, and treatment modalities.
†Adjusted for age group, sex, region of hospitals, comorbidities (Diabetes, chronic lung disease, and major neurologic diseases), Charlson comorbidity index, treatment modalities, and presence of severe infection. ARB denotes Angiotensin receptor blocker, ACEI angiotensin-converting enzyme inhibitor, aOR adjusted odds ratio, and CI confidence interval

**Supplementary Table S4.**
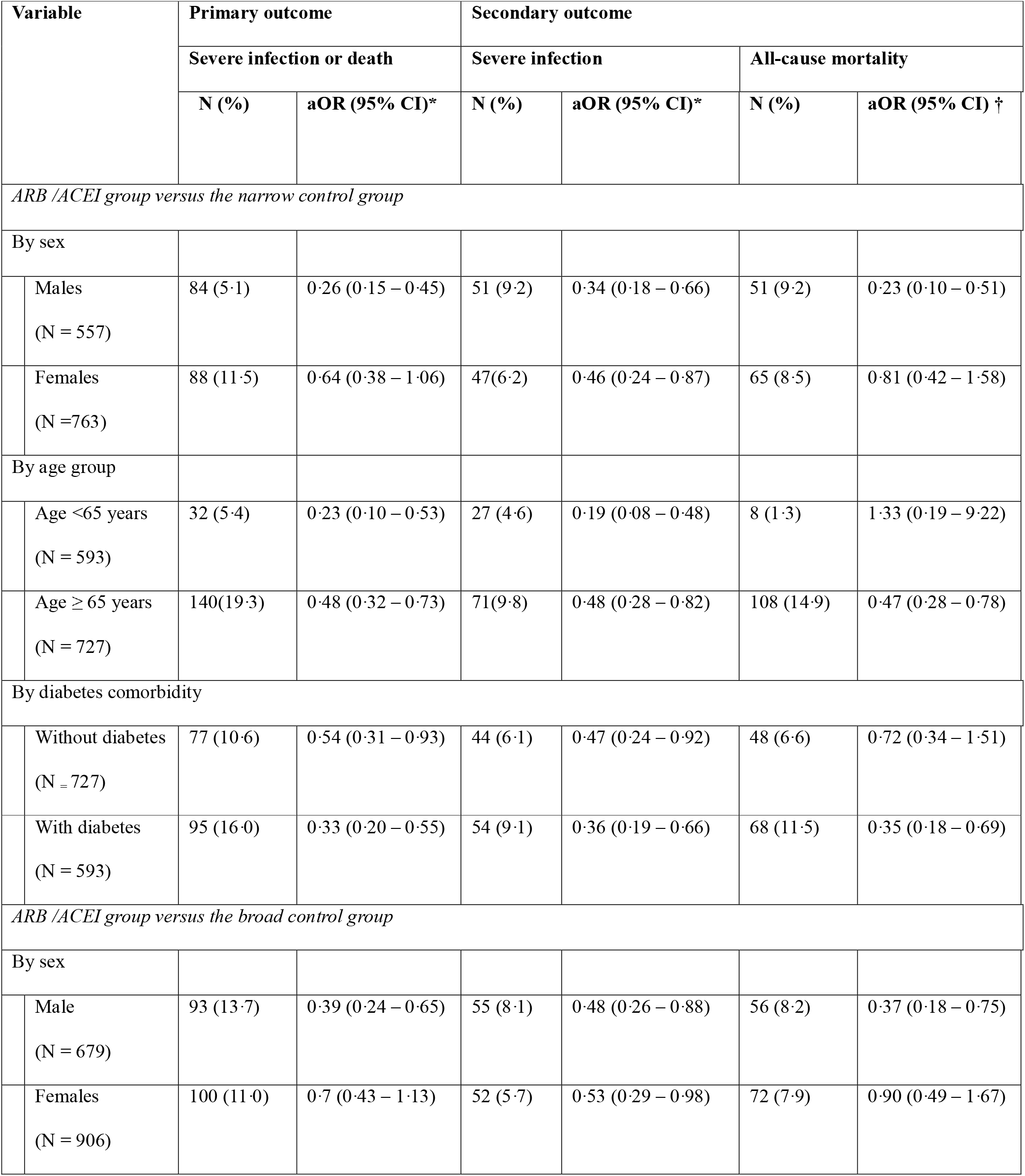
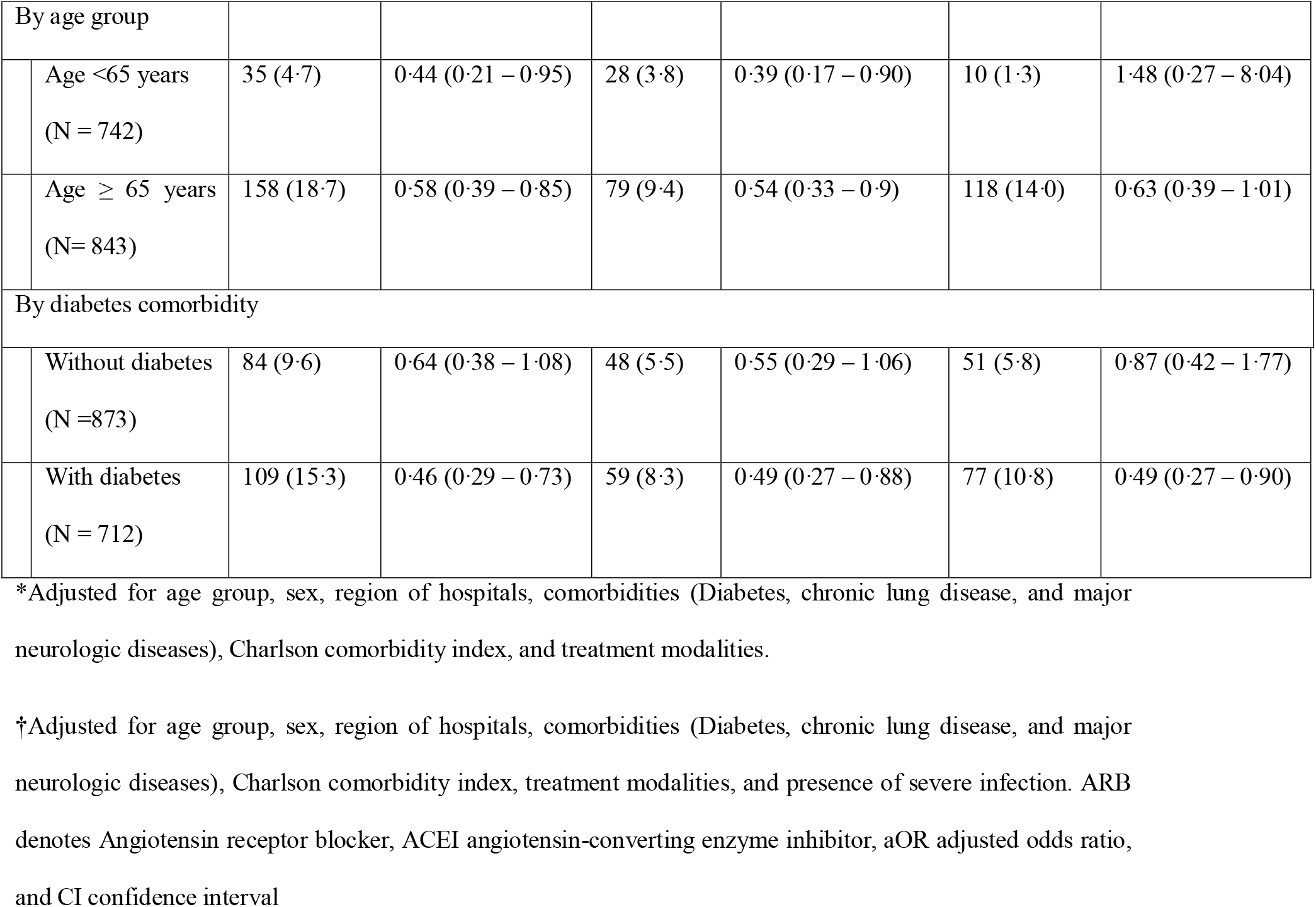
Subgroup analysis according to sex, age group, and diabetes comorbidity.

